# A Computational Growth Framework for Biological Tissues: Application to Growth of Aortic Root Aneurysm Repaired by the V-shape Surgery

**DOI:** 10.1101/2021.09.30.21264318

**Authors:** Hai Dong, Minliang Liu, Tongran Qin, Liang Liang, Bulat Ziganshin, Hesham Ellauzi, Mohammad Zafar, Sophie Jang, John Elefteriades, Wei Sun, Rudolph L. Gleason

**Author notes:** **For correspondence:** Rudolph L. Gleason Jr, Ph.D., The Wallace H. Coulter Department of Biomedical Engineering, Georgia Institute of Technology and Emory University, Technology Enterprise Park, Room 204, 387 Technology Circle, Atlanta, GA 30313-2412.

## Abstract

Ascending aortic aneurysms (AsAA) often include the dilatation of sinotubular junction (STJ) which usually leads to aortic insufficiency. The novel surgery of the V-shape resection of the noncoronary sinus, for treatment of AsAA with root ectasia, has been shown to be a simpler procedure compared to traditional surgeries. Our previous study showed that the repaired aortic root aneurysms grew after the surgery. In this study, we developed a novel computational growth framework to model the growth of the aortic root repaired by the V-shape surgery. Specifically, the unified-fiber-distribution (UFD) model was applied to describe the hyperelastic deformation of the aortic tissue. A novel kinematic growth evolution law was proposed based on existing observations that the growth rate is linearly dependent on the wall stress. Moreover, we also obtained patient-specific geometries of the repaired aortic root post-surgery at two follow-up time points (Post1 and Post2) for 5 patients, based on clinical CT images. The novel computational growth framework was implemented into the Abaqus UMAT user subroutine and applied to model the growth of the aortic root from Post1 to Post2. Patient-specific growth parameters were obtained by an optimization procedure. The predicted geometry and stress of the aortic root at Post2 agree well with the *in vivo* results. The novel computational growth framework and the optimized growth parameters could be applied to predict the growth of repaired aortic root aneurysms for new patients and to optimize repair strategies for AsAA.

## 1. Introduction

Aortic aneurysms are critical conditions that frequently require surgical intervention to prevent dissection, rupture, and death [1-4]. Ascending aortic aneurysms (AsAA) often include the dilatation of sinotubular junction (STJ) and extend proximally into the the aortic root, which usually leads to aortic insufficiency [5, 6]. Recently, Elefteriades et al. [7] reported a simpler V-shape resection technique of the noncoronary sinus intended for infirm or elderly patients with AsAA and root ectasia. This simple, quick procedure addresses the aortic root dilatation without the complexities of full aortic root surgery with coronary button reimplantation [7]. In our previous study, we showed [8] that there is significant reduction in the wall stress of the aortic root repaired by the V-shape surgery. The results also indicated that the aortic root could grow after the V-shape surgery. It will be beneficial to model the growth of the aortic root aneurysm after surgery for growth prediction of new patients.

A number of studies have investigated the growth and remodeling (G&R) of aortic aneurysms. For instance, Lin et al. [9] studied the formation and dilatation of abdominal aortic aneurysms with a mixture theory of growth and remodeling. Mousavi et al. [10] predicted the aneurysm growth and remodeling in the ascending thoracic aorta using the homogenized constrained mixture model. Ghavamian et al. [11] studied of growth and remodeling in ascending thoracic aortic aneurysms considering variations of smooth muscle cell basal tone. Farsad et al. [12] investigated the effect of the spine constraint for progression of the abdominal aortic aneurysms using a membrane G&R model. However, few existing studies have modelled the growth and remodeling of aortic aneurysms with follow-up clinical images for patient-specific geometries.

The constrained mixture model [13] and the kinematic/finite growth model [14] are two leading approaches for growth and remodeling of soft tissues. The constrained mixture model, proposed by Humphrey and Rajagopal [13], considers the kinetics of the production and removal of individual constituents and has been widely used to model the growth and remodeling of soft tissues including myocardial hypertrophy [15], tendon healing [16], and growing tumor [17]. The kinematic/finite growth model, developed by Rodriguez et al. [14], separates the total deformation into an elastic part and a growth part, and has also been extensively applied in growth and remodeling of soft tissues. For instance, Eskandari et al. [18] studied the patient-specific airway wall remodeling in chronic lung disease by the kinematic growth model. Soleimani et al. [19] developed a stress-induced anisotropic growth model driven by nutrient diffusion based on the finite/kinematic growth approach. The kinematic growth approach is appropriate for studies that do not quantify changes in the content and organization of individual constituents, as is often the case in studies that rely solely on non-invasive measurements.

In this paper, we developed a novel computational framework with the kinematic growth approach for the growth of the aortic root following repaired by the V-shape surgery. The unified-fiber-distribution (UFD) model [20] was applied to describe the hyperelastic deformation of the aortic tissue. A novel kinematic growth evolution law was proposed based on existing observations that the growth rate is approximately linearly dependent on the principal components of in-plane wall stress. The novel computational growth framework was implemented into the Abaqus UMAT user subroutine and applied to model the growth of the aortic root aneurysm repaired by the V-shape surgery, based on patient-specific follow-up geometries reconstructed from *in vivo* CT images. Patient-specific growth parameters were obtained from the first four patient data sets by an optimization procedure. The growth framework, using the average growth parameters from the first four patients, were compared to the follow-up geometries of the fifth patient to assess the predictive capability of the model and fitted parameters. The wall stress of the aortic root was also investigated.

## 2. Methods

### 2.1 Construction of Patient-Specific Finite Element Model

Clinical CT images of the aortic root at two follow-up time points post-surgery were obtained for 5 patients (P1-P5) who underwent the surgery of the V-shape resection of the noncoronary sinus [7], at Yale-New Haven Hospital, New Haven, Connecticut. The CT image of the first time point (Post1) was obtained within 4 months after the surgery. The time interval between the second time point (Post2) and Post1 ranged from 20 months to 52 months. The patient demographic information is provided in Table 1. The in-plane spatial resolution of the CT images was between 0.37 × 0.37 mm and 0.96 × 0.96 mm, with the slice thickness ranging from 0.30 to 1.00 mm. The details of the CT images resolution for each patient can be found in the Appendix. All human subjects in this study provided written and informed consent, and IRB approvals were obtained from Georgia Tech Institutional Review Board (#H20373, 10/13/2020) and Yale University Human Investigation Committee (#0109012617, 06/29/2020).

**Table 1:** Patient information: Gender, systolic pressure, diastolic pressure, and time interval (ΔT_12_) between the two follow-up time points post-surgery (Post1 and Post2).

| Patient ID | Gender | Systolic Pressure (mmHg) | Diastolic Pressure (mmHg) | $\Delta T_{12}$ (month) |
| --- | --- | --- | --- | --- |
| P1 | F | 123 | 92 | 20 |
| P2 | M | 140 | 79 | 43 |
| P3 | M | 125 | 82 | 24 |
| P4 | M | 120 | 62 | 52 |
| P5 | M | 120 | 80 | 22 |

Based on the CT images (e.g., Fig. 1a), the three-dimensional (3D) surface geometry (Fig. 1b) of the aortic root was reconstructed using 3D Slicer (www.slicer.org). The surface geometry was exported out from the 3D Slicer and imported into Altair HyperMesh 2017 (Altair Engineering) to generate the finite element mesh for the aortic root (Fig. 2c). We first created 4-node quadrilateral shell elements (S4) with element size of about 1.5 mm by 1.5 mm based on the surface geometry. Four layers of eight-node linear brick elements (C3D8) were created by offsetting the S4 elements [21, 22]. A total thickness estimate of 1.5 mm was applied to the aortic wall following Liu et al. [23], since the accurate thickness of the aortic wall is not available from the CT images. The finite element mesh of the aortic root of Post1 and Post2 for the 5 patients were obtained. Analysis of mesh size independence was performed.

**Fig 1.**
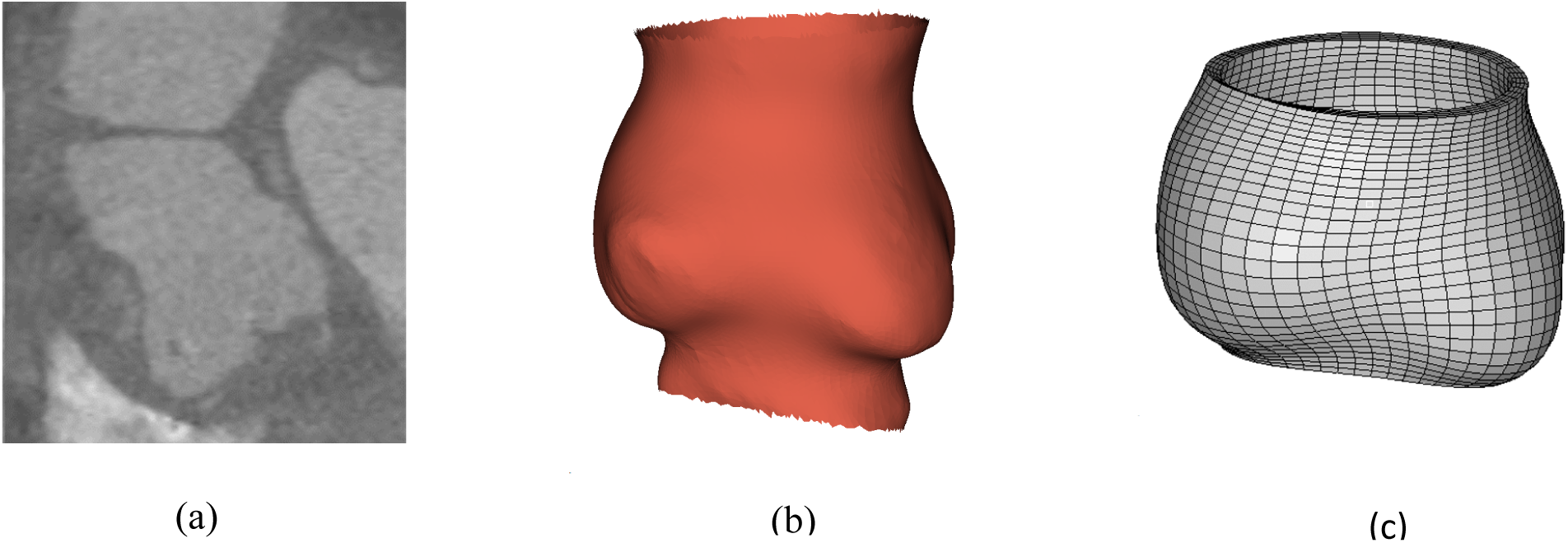
(a) CT image of the aortic root of a representative patient (P1) post-surgery; (b) 3D surface geometry of the aortic root reconstructed based on CT in (a) using 3D Slicer (www.slicer.org); (c) Finite element mesh generated in HyperMesh 2017 (Altair Engineering) based on the geometry in (b) with trimmed boundaries.

**Fig 2.**
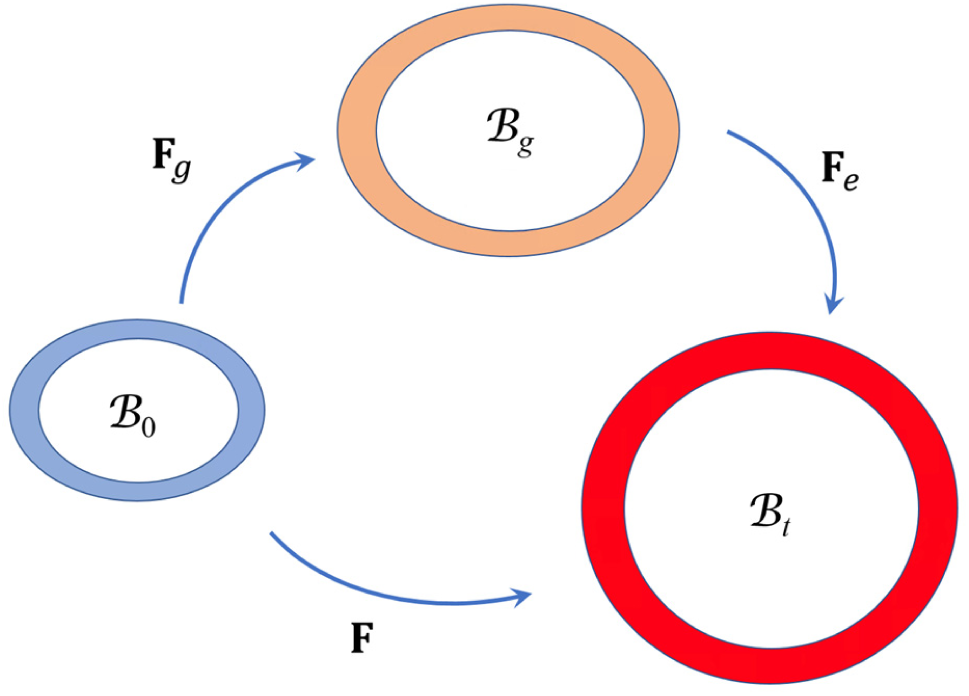
Schematic of the multiplicative decomposition of the total deformation gradient **F** into an elastic part **F**_e_ and a growth part **F**_g_. *ℬ*_0_ represents the reference configuration, *ℬ*_*g*_ is the growth configuration and *ℬ*_*t*_ denotes the current configuration under inner pressure loading.

### 2.2 Growth Framework based on the Unified-Fiber-Distribution Model

Following Rodrigue et al. [14], we assumed that the total deformation gradient of the tissue can be decomposed multiplicatively into an elastic part and a growth part (Fig. 2),

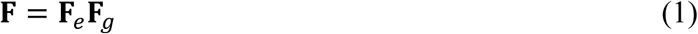

where **F**_*g*_ is the deformation gradient from the original reference state to the current reference state due to tissue growth, and **F**_*e*_ is the elastic deformation gradient from the current reference state to the current loaded state due to the mechanical loading. The constitutive model for the relation between the elastic deformation **F**_*e*_ and the loading was described by our recently developed unified-fiber-distribution (UFD) model [20] which considers the fiber distribution as a whole distribution without separating it into multiple fiber families. The evolution of the growth deformation **F**_*g*_ was described by a newly proposed growth evolution law based on the assumption that the tissue growth was governed by the in-plane principal components of Cauchy stress.

#### 2.2.1 Unified-Fiber-Distribution (UFD) Hyperealstic Model

Aortic walls are usually composed of three layers, the intima, media and adventitia layers [24]. The tissue of each layer is comprised of networks of collagen fibers embedded in a ground matrix and can be regarded as fiber reinforced composites, of which the mechanical properties are anisotropic [25-27], similar to engineering fiber-reinforced composites [28-32]. Schriefl at al. [33] showed that there are significant dispersions for the collagen fiber distributions in the three layers of human arteries, and each layer may contain multiple fiber families. Their study [33] also revealed that the number of fiber family may vary from different positions for the same kind of tissue, such as there are two fiber families in the media of human descending thoracic aorta and abdominal aorta, while there is only one fiber family in the media of common iliac artery. Previously, we developed a unified-fiber-distribution (UFD) model [20] which considers the fibers as a unified distribution and thus the number of fiber families is not necessary as it is in the widely used Gasser–Ogden–Holzapfel (GOH) model [34]. The later usually reduces a general fiber distribution into two fiber families, which may be not consistent with the real fiber distribution of aortic tissues. Here, the UFD model was applied to characterize the hyperelastic deformation of the aortic tissues.

The total strain energy function can be decomposed additively into an isochoric part and a volumetric part

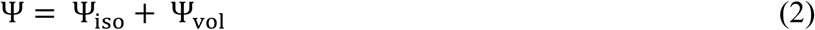

where the volumetric part is given by 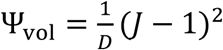 with *D* to be a material constant which is used to enforce incompressibility of the tissue, based on the assumption that the aortic tissue is approximately incompressible. The isochoric part can be further expressed as

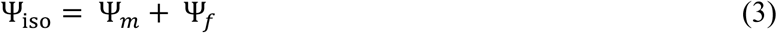

where Ψ_*m*_ is the strain energy of the matrix contribution given by the neo-Hookean model [35-39]

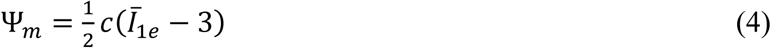

where *c* is the initial shear modulus of the matrix, 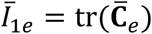 is the first invariant of 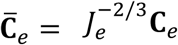, with 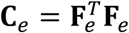 denoting the right Cauchy–Green tensor and *J*_*e*_ = det**F**_*e*_ > 0 is the determinant of the deformation gradient **F**_*e*_. The Ψ_*f*_ is the strain energy of the fiber contribution which distinguishes the UFD model from other models, given by [20]

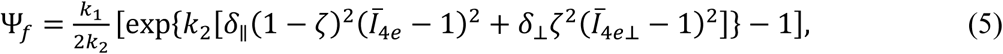

where 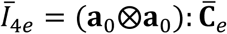, and 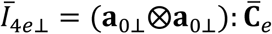 with **a**_0_ and **a**_0⊥_ denoting the unit vectors in the circumferential (mean-fiber) direction and the axial (cross-mean-fiber) direction, respectively. The parameters *k*_1_, *k*_2_ and ζ are fiber related material parameters with *k*_1_ representing the initial stiffness, *k*_2_ representing the stiffening effect under stretch, and ζ is a scalar characterizing the fiber distribution in an integral sense. The role of *δ*_‖_ and *δ*_⊥_ is to exclude the contribution of the compressed fibers, given by the Heaviside step function

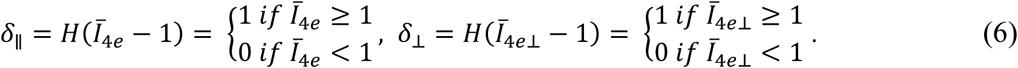

The material parameters of the UFD model applied in this study are summarized in Table 2.

**Table 2.**
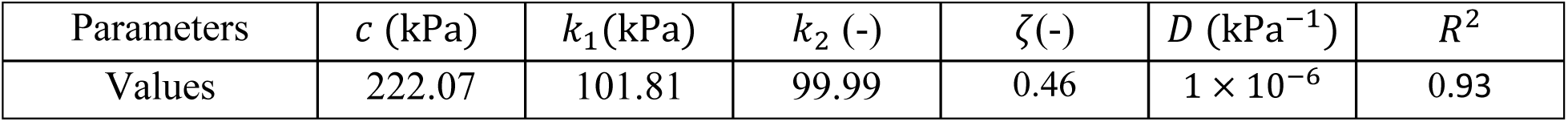
The parameters used in the UFD model. The *R*^2^ describes the goodness of the fit based on the experiments [40].

#### 2.2.2 Growth Evolution Law

We assumed the growth deformation **F**_*g*_ in Eq. (1) can be expressed as

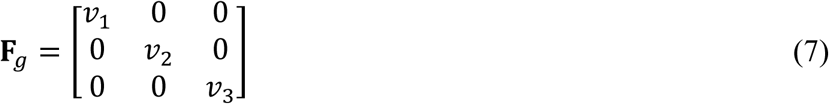

where 𝒱_1_, 𝒱_2_ and 𝒱_3_ are the growth stretch multipliers. The directions {1, 2, 3} represent the principal directions of the local Cauchy stress. For a cylindrical tube under inner pressure loading, the directions {1, 2, 3} represents the circumferential, axial and radial directions, respectively; however, in a non-cylindrical aortic root, these directions may not align with these directions.

It has been shown that the aortic wall stress is highly related to dilation of aortic aneurysms. For example, Shang et al. [41] obtained a linear relation (*R*^2^ = 0.81) between the peak wall stress and the expansion rate of aortic aneurysms without surgery reparation. More recently, our group [8] obtained a linear relation (*R*^2^ = 0.92) between the mean max-principal stress and the dilation rate of aortic root aneurysm after surgery reparation. Bearing these in mind, we proposed that the evolution of the growth rate 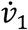 is linearly dependent on σ _1_, the max-principal component of the Cauchy stress, and the growth rate of 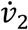 linearly dependent on the σ _2_, the mid-principal component of the Cauchy stress. As mentioned in **Sec. 2.1**, the thickness of the aortic wall is not available and thus we assume that the thickness remains between Post1 and Post2. The growth evolution laws can be summarized as

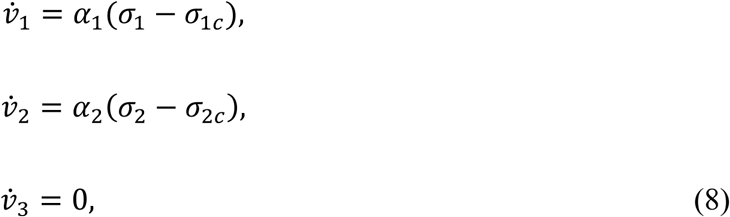

where {*α*_*i*_, *σ*_*ic*_} (*i* = 1, 2) are growth parameters. The parameters *σ*_*ic*_ (i = 1, 2) > 0 can be regarded as the critical trigger stress for the growth, i.e. the growth is triggered when the stress *σ*_*i*_ is greater than *σ*_*ie*_ and there is no growth when the stress *σ*_*i*_ is less than *σ*_*ie*_. The true stress subtracted by the critical trigger stress yields the pure growth stress which contribute to the growth.

The UFD model and growth evolution law were implemented into the UMAT user-defined material subroutine of the Abaqus FEA 2020 (SIMULIA, Providence, RI). A numerical approximation scheme [42, 43] was applied to calculated the tangent modulus tensor. The implemented model was applied to simulate the growth of the aortic root from Post1 to Post2. The growth parameters {*α*_*i*_, *σ*_*ie*_} for each patient were obtained by an optimization procedure based on the aortic root geometries at Post1 and Post2, which will be explained detail in the next section.

### 2.3 Growth Simulation and Optimization of Growth Parameters

Figure 3 shows the flow chart of the growth simulation and optimization of the growth parameters. We simulated the growth of the aortic root for each patient starting from the geometry of the aortic root of Post1 (Fig. 3a). Since the geometry of the aortic root was reconstructed based on the CT scans of the *in vivo* configuration under systolic pressure, applying the physiological systolic pressure directly on the in vivo geometry would obtain inaccurate strain and stress. Instead, the FE simulation should start from the zero-pressured geometry. We used the improved backward displacement method [44] to obtain the zero-pressured geometry of the aortic root of Post1 (Fig. 3b).

**Fig 3.**
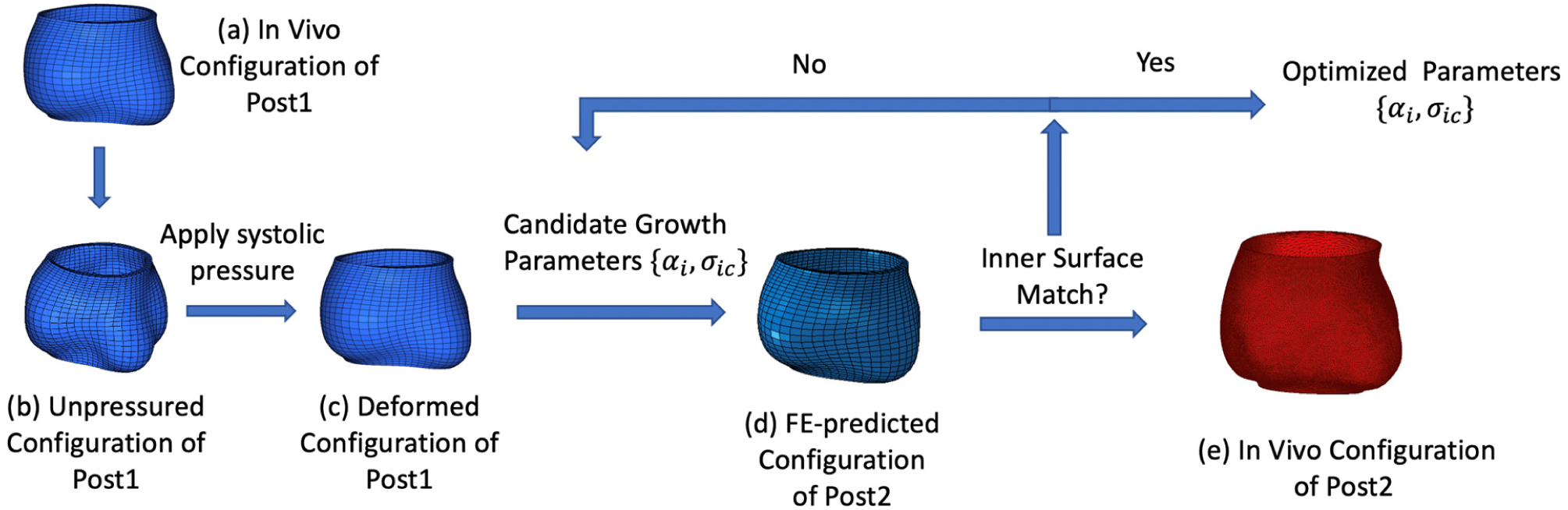
Flow chart of the growth simulation and optimization of growth parameters.

Based on the zero-pressured geometry of the aortic root of Post1, the finite element (FE) simulation of the growth was performed in two steps. In the first step, the patient-specific systolic pressure was applied to the inner surface of the zero-pressured geometry of the aortic root of Post1 to obtain the stress and strain fields (Fig. 3c). During the first step, there is no growth for the aortic tissue. In the second step, the growth process was activated with a group of candidate growth parameters and the growth stretch was calculated following the growth laws in Eqs. (8) and (9), based on the stress field obtained from the first step. The growth process was stopped when the growth period reached the time-point Post2 and the FE-predicted configuration of the aortic root at Post2 was obtained (Fig. 2d). The growth parameters {*α*_*i*_, σ_ie_} was tuned until the inner surface of the FE-predicted configuration of Post2 matched the *in vivo* inner surface (Fig. 3e) of Post2 for the aortic root.

Specifically, the method of least squares was applied to match the FE-predicted and *in vivo* root inner surface of Post2. We define the node-to-surface distance between a mesh node **X** of the predicted root inner surface (extracted from Fig. 3d) and the triangle mesh Ω of the *in vivo* root inner surface of Post2 (Fig. 3e) as

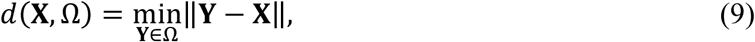

where **X** is the coordinate vector of the node, **Y** is the coordinate vector of a node on the mesh surface Ω, and the operator ‖**Z**‖ represents the 3D Euclidean norm of an arbitrary vector **Z**. The value of *d*(**X**, Ω) gradually converges to a specific value with the increasing mesh density (decreasing mesh size) of Ω. In this study, the distance converging mesh size of Ω was around 0.3 mm. The growth parameters {*α*_*i*_, *σ*_*ie*_} was optimized by minimizing the sum of the squares of the node-to-surface distance between the FE-predicted and *in vivo* root inner surface.

We implemented the optimization procedure based on the function *lsqnonlin* in MATLAB 2018b (MathWorks, Natick, MA). The FE simulations were performed by Abaqus FEA 2020 (SIMULIA, Providence, RI) using a cluster server (Partnership for an Advanced Computing Environment (PACE) at the Georgia Institute of Technology). The local optimization MATLAB program needs to communicate with the PACE server, including uploading files to, locally submitting jobs to, and downloading result files from the server. The uploading and downloading of files were realized by the python version of SFTP (Pysftp, https://pypi.org/project/pysftp/), and the paramiko (http://www.paramiko.org/) was used to locally submitting jobs to the server.

The root inner surface of Post1 before growth is enclosed inside of the root inner surface of Post2 (Fig. 4). Part of the root surface of Post1 could grow outside of the root surface of Post2 while other parts may still remain enclosed inside by the surface of Post2 after the growth. In order to reflect the relative position of a node on the root inner surface of Post1 after growth to the root inner surface of Post2, we defined another distance parameter with a sign as

**Fig 4.**
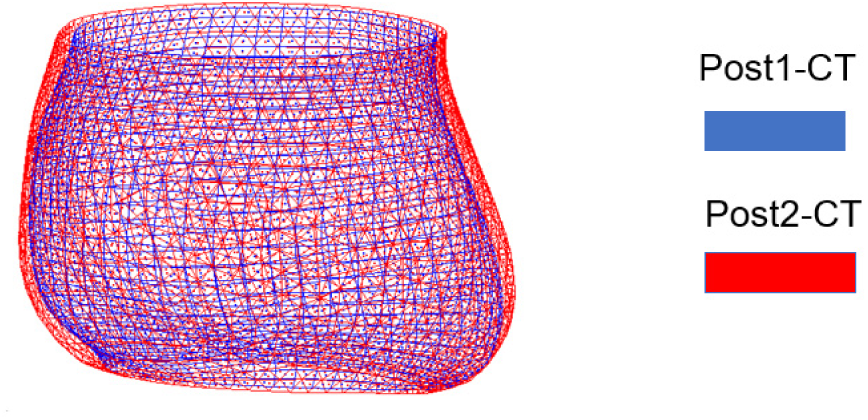
*In vivo* geometries of the aortic root for a representative patient (P1) at two follow-up time points post-surgery (Post1 and Post2), reconstructed from the clinical CT images.

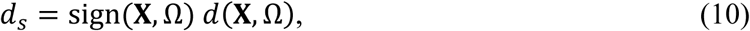

Where *d*(**X**, Ω) is given in Eq. (9), with sign(**X**, Ω) = 1 when **X** located inside the surface Ω and sign(**X**, Ω) = -1 when **X** located outside the surface Ω.

## 3. Results

### 3.1 Optimization Procedure and Optimized Growth Parameters

Figure 5a shows the comparison between the FE-predicted geometry of Post2 (growth from Post1, Post1-Growth) at the first iteration and Post2 from CT, for the aortic root of a representative patient (P1). The corresponding *in vivo* geometries of Post1 and Post2 from CT have been shown in Fig. 4. The results in Figure 5a indicate that the growth from Post1 is too much, with the mean node-to-surface distance between Post1-Growth and Post2-CT to be -0.62 mm, the negative sign of the distance indicating that the overall surface of Post1 has grown outside the *in vivo* surface of Post 2. The candidate growth parameters were then tuned and the mean node-to-surface distance between Post1-Growth and Post2-CT reduced (in the sense of absolute value) gradually to the final value of +0.09 mm (Fig. 5b). Similar results were obtained for patients P2-P4 during the optimization process.

**Fig 5.**
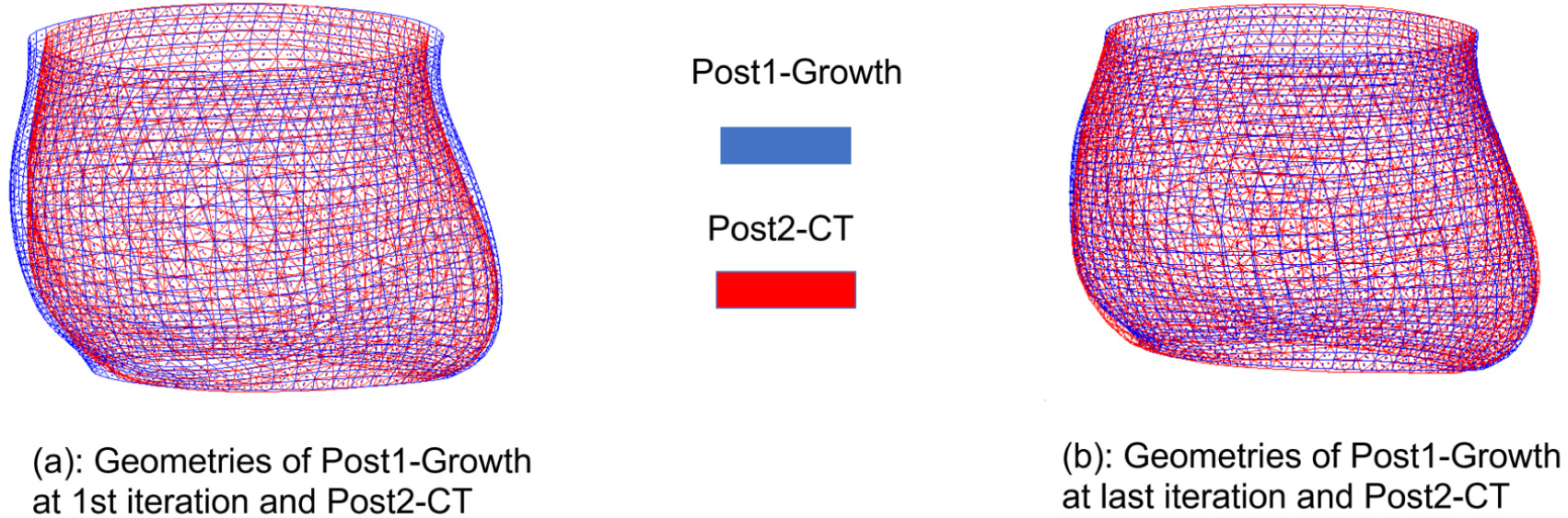
(a) Comparison between FE-predicted geometry of Post2 (growth from Post1, Post1-Growth) at the first iteration and *in vivo* geometry of Post2 from CT (Post2-CT), for a representative patient (P1); (b) comparison between FE-predicted geometry of Post2 (Post1-Growth) at the last iteration and Post2-CT.

The mean node-to-surface distance between the *in vivo* inner surface of Post1 and Post2 for the aortic root of the 4 patients (P1-P4) ranges from +0.79 mm to +1.38 mm (Table 3). The mean node-to-surface distance between the FE-predicted inner surface of Post2 (growth from Post1) with the optimized growth parameters (Table 4) and the *in vivo* geometry Post2, ranging from -0.04 to 0.09 mm. For all the patients, the value of *α*_1_ is greater than *α*_2_, and the value of *σ*_1*c*_ is less than *σ*_2*c*_.

**Table 3:** Mean+standard deviation (mean+std) of the node-to-surface distance between the *in vivo* geometries of Post1 and Post2 from the CT images (Post1-CT & Post2-CT) for the aortic root of the 4 patients (P1-P4), and the node-to-surface distance between predicted geometry of Post2 based on growth from Post1 (Post1-Growth) and *in vivo* geometry of Post2-CT. A negative mean distance (e.g., P3 and P4) between Post1-Growth and Post2-CT represents the overall root surface of Post1-Growth grows outside of the root surface of Post2-CT. The corresponding optimized growth parameters are listed in Table 4.

| Distance (mm) | Post1-CT& Post2-CT (mean±std) | Post1-Growth & Post2-CT (mean±std) |
| --- | --- | --- |
| P1 (P13) | 0.79 ± 0.40 | 0.09 ± 0.39 |
| P2 (P07) | 1.11 ± 0.52 | 0.08 ± 0.63 |
| P3 (P03) | 1.38 ± 0.77 | -0.02 ± 0.71 |
| P4 (P16) | 1.22 ± 0.97 | -0.04 ± 0.93 |

**Table 4:** Optimized growth parameters in Eq. (9) for the aortic root of the 4 patients (P1-P4), corresponding to the predicted results of Post2 based on the growth from Post1 (Post1-Growth) in Table 3.

| | $\alpha_1$ (1/[MPa · Mon]) | $\alpha_2$(1/[MPa · Mon]) | $\sigma_{1c}$ (kPa) | $\sigma_{2c}$(kPa) |
| --- | --- | --- | --- | --- |
| P1 (P13) | 0.0119 | 0.0045 | 82.5 | 290.0 |
| P2 (P07) | 0.0048 | 0.0015 | 157.1 | 240.7 |
| P3 (P03) | 0.0114 | 0.0065 | 38.0 | 216.6 |
| P4 (P16) | 0.0025 | 0.0020 | 23.2 | 168.0 |
| Average | 0.0077 | 0.0036 | 75.2 | 228.8 |

### 3.2 Prediction of P5 based on Parameters of P1-P4

The aortic root of P5 grows mildly from Post1 to Post2 with the mean and std node-to-surface distance to be 0.85 + 0.51 mm. The comparison between the predicted geometry of Post2 (Post1-Growth), based on the average value of the optimized growth parameters of P1-P4 in Table 4, and Post2-CT suggests a good match between the predicted and *in vivo* geometries of Post2, with the mean node-to-surface distance between Post1-Growth and Post2-CT to be -0.17 + 0.58 mm (Fig. 6).

**Fig 6.**
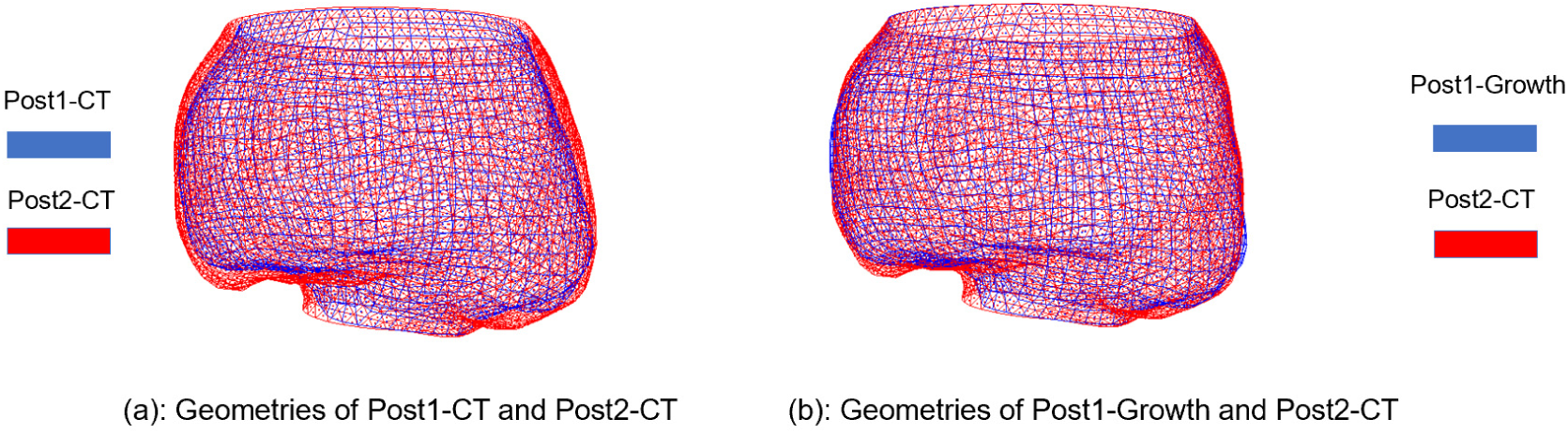
(a) Comparison between the *in vivo* geometries of Post1 from CT (Post1-CT) and Post2 from CT (Post2-CT) for the aortic root of P5; (b) comparison between the predicted geometry of Post2 (growth from Post1, Post1-Growth) and Post2-CT.

### 3.3 Aortic Root Wall Stress

We also obtained the wall stress of the aortic root of Post2 based on the predicted and *in vivo* geometries. Figure 7 showed the max-principal stress field of the aortic root of Post2 for a representative patient (P1), which suggests that the overall stress distribution follows similar pattern for results based the predicted (Fig. 7a) and *in vivo* (Fig. 7b) geometries. Table 5 presented the mean and peak values of the max-principal stress of Post2 based on the predicted and *in vivo* geometries for the five patients (P1-P5). The stress based on predicted geometry agrees well with that based on the *in vivo* geometry, with an average difference of -3.83% for the mean value and - 4.12% average difference for the peak value.

**Table 5:** Mean and peak values of the max-principal stress of Post2 based on the predicted and *in vivo* geometries for the five patients (P1-P5).

|  | Mean Stress (kPa) |  |  | Peak Stress (kPa) |  |  |
| --- | --- | --- | --- | --- | --- | --- |
|  | Predicted | In Vivo | Difference | Predicted | In Vivo | Difference |
| P1-Post2 | 163.01 | 164.88 | -1.13% | 262.11 | 266.56 | -1.67% |
| P2-Post2 | 202.75 | 224.04 | -9.50% | 287.56 | 315.68 | -8.91% |
| P3-Post2 | 181.82 | 184.38 | -1.39% | 321.14 | 359.22 | -10.60% |
| P4-Post2 | 182.63 | 202.5 | -9.81% | 278.97 | 293.77 | -5.04% |
| P5-Post2 | 157.52 | 148.11 | 6.35% | 384.35 | 359.99 | 6.77% |
| Average | 177.546 | 184.782 | -3.92% | 306.826 | 319.044 | -3.83% |

**Fig 7.**
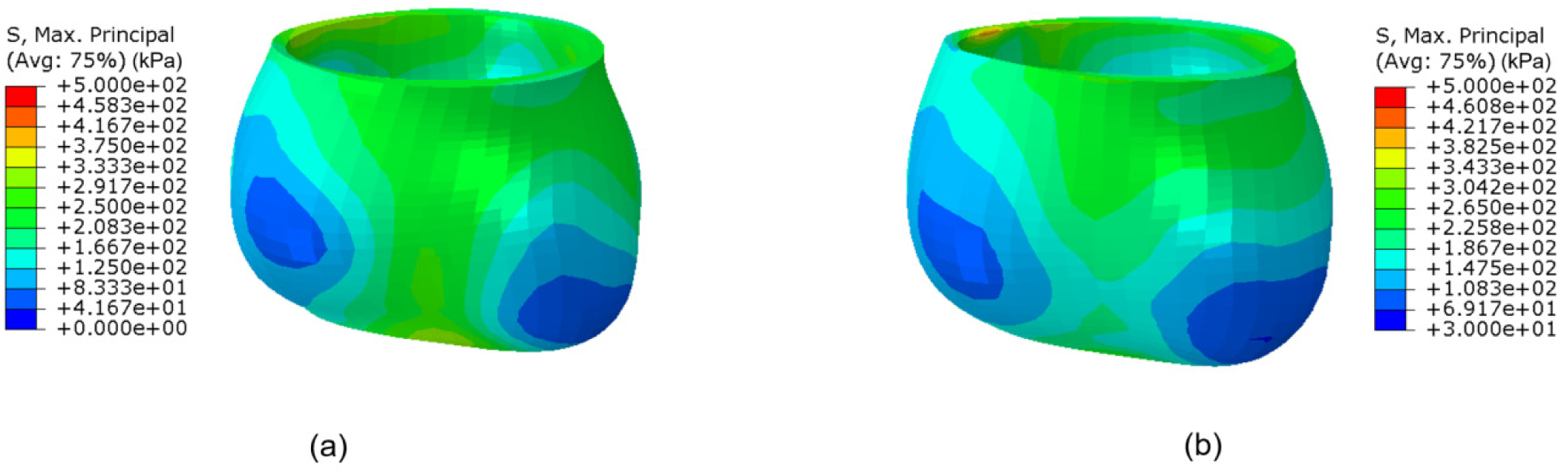
Max-principal stress field of the aortic root of Post2 for a representative patient (P1): (a) results based on the predicted geometry; (b) results based on *in vivo* geometry. The stress distributions follows similar pattern.

## 4. Discussions

We developed a novel computational growth framework for biological tissues based on the unified-fiber-distribution (UFD) model and kinematic growth approach, in which growth is governed by the two in-plane principal components of the Cauchy stress. The UFD model [20] considers the fiber distribution in aortic tissues as a unified distribution which is expected to be more physically consistent, with the real fiber distribution in aortic tissues, than separating the fiber distributions into two/several fiber families. For the kinematic growth evolution law, four growth parameters {*α*_1_, *α*_2_, *σ*_1*e*_, *σ*_2*e*_} were used in Eq. (9). The parameters {*α*_1_, *α*_2_} control the growth rate under specific stress conditions, and the parameters {*σ*_1*e*_, *σ*_2*e*_} represent the critical trigger value of the Cauchy stress that could induce the growth. For instance, for the optimized parameters of P1 in Table 4, the value of *σ*_1*e*_ =82.5 kPa indicates that there will be no growth in 1 direction when *σ*_1_ is less than 82.5 kPa, and the value of *α*_1_ = 0.0119 [MPa · Mon]^1^ suggests that the growth stretch will be 0.119% per month under the pure growth stress value of 100 kPa (Cauchy stress subtracting the growth trigger stress).

As mentioned before in Sec. 2.2.2, the principal stresses *σ*_1_ and *σ*_2_ are along the circumferential and axial directions, respectively, for a perfect cylindrical tube under inner pressure loading. The geometry of the aortic root (e.g, Fig. 1c) is a cylinder-like tube, and the principal stresses *σ*_1_ and *σ*_2_ in the aortic root are approximately along the circumferential and axial directions, respectively. The value of the optimized growth parameter *α*_1_ is greater than that of *α*_2_ for all patients (Table 4), which suggests that the growth rate in the circumferential direction is larger than that in axial direction under the same pure growth stress. The optimized value of *σ*_1*c*_ is smaller than that of *σ*_2*c*_ for all patients indicates that the growth in the circumferential direction is easier to be triggered than that in the axial direction.

We used the growth parameters of P1-P4 to predict the growth of the aortic root of P5, which illustrated the possibility of predicting growth of new patients based on parameters of existing patients. It is possible that predictions of growth for other new patients based on the database of P1-P4 are not as accurate as that of P5, since the current database contains a rather limited number of patients. The database of the growth parameters will be expanded with follow-up CT images of more patients being accumulated. Vande Geest et al. [45] showed that the patient-specific material property of aortic wall (e.g., wall strength) is possible to be estimated from existing database depending on clinical factors such as age, gender, family history, etc. The patient-specific growth parameters may also depend on these clinical factors. The parameters for new patients estimated based on the approach of age-gender-match from a larger parameter database may produce more accurate prediction of the growth.

Long-time consuming is one of the most distinguishing features for optimization problems, since the optimization process usually requires multiple iterations in order to obtain the optimized solution. The optimization of the growth parameters in this study is even more complex because the finite element (FE) simulation result is needed for each iteration during the optimization procedure. We used a combined technique which includes the local PC for optimization procedure and the cluster server to perform the FE simulation. Comparing to running all the computations with a local PC, such kind of combined technique could reduce the costing time in two way: 1) the FE simulation performed on the cluster server is significantly faster than that on a local PC; 2) the optimization for different patients could be performed simultaneously since a cluster server has less limited (or unlimited) CPU resources.

There are several limitations of this study. First, the *in vivo* thickness of the aortic root was not available from the CT images, and thus the potential growth in the radial direction from Post1 to Post2 was not considered in our simulation. As shown by others, the time rate of change of radial versus in-plane growth can be an important factor in the growth rate of aneurysms [46-48] and should be a topic of future work. *In vivo* thickness of the aortic wall may be obtained based on the method in Elefteriades et al. [49] when both contrast and non-contrast CT scans are available. Second, although the patient-specific growth parameters were obtained from the optimization procedure, only one set of hyperelastic material properties derived from experimental *ex vivo* data of human aortic aneurysm tissue were used for all the patients in this study. Patient-specific elastic material properties may be obtained by inverse methods [50, 51] when two-phase CT images under different pressure loadings are available. Finally, we only have five patients with CT images available at two follow-up time points for the growth simulation. Future studies should include larger cohort of patients with more CT data being accumulated.

## 5. Conclusions

In this study, we developed a novel computational growth framework for biological tissues and applied it to simulate the growth of the aortic root aneurysm repaired by the V-shape surgery. Particularly, the unified-fiber-distribution (UFD) model was applied to describe the hyerelastic deformation of the aortic tissue. A novel kinematic growth evolution law was proposed based on existing observations that the growth rate is linearly dependent on the wall stress. Moreover, we also obtained patient-specific geometries of the aortic root aneurysm post-surgery at two follow-up time points (Post1 and Post2) for 5 patients, based on clinical CT images. The novel computational growth framework was implemented into the UMAT user subroutine of the Abaqus FEA, and applied to model the growth of the aortic root from Post1 to Post2. Patient-specific growth parameters were obtained by an optimization procedure. The predicted geometry and stress of the aortic root at Post2 agree well with the *in vivo* results. The novel computational growth framework and the optimized growth parameters could be applied to predict the growth of repaired aortic root aneurysms for new patients.

## Data Availability

All the data have been presented in the manuscript

## Acknowledgment

H. Dong thanks lab members Courtney Huynh, Samuel Li, and Kristen Riess for assistance in image segmentation, mesh creation and finite element simulation. This study is in part supported by American Heart Association (AHA) 18TPA34230083.

## Appendix: Resolution of CT images

The in-plane spatial resolution of the CT images of the five patients (P1-P5) is between 0.37 × 0.37 mm and 0.96 × 0.96 mm, with the slice thickness ranging from 0.30 to 1.00 mm. The details of the CT images resolution for each patient were listed in Table A1.

**Table A1:** In-plane resolution and slice thickness of CT images of Post1 and Post2 for aortic root of P1-P5.

| Patient | In-plane resolution of Post1 (mm) | Slice thickness of Post1 (mm) | In-plane resolution of Post2 (mm) | Slice thickness of Post2 (mm) |
| --- | --- | --- | --- | --- |
| P1 | $0.49 \times 0.49$ | 0.63 | $0.71 \times 0.71$ | 1.00 |
| P2 | $0.63 \times 0.63$ | 0.63 | $0.96 \times 0.96$ | 0.63 |
| P3 | $0.69 \times 0.69$ | 0.30 | $0.69 \times 0.69$ | 0.30 |
| P4 | $0.78 \times 0.78$ | 0.63 | $0.74 \times 0.74$ | 0.63 |
| P5 | $0.67 \times 0.67$ | 0.30 | $0.37 \times 0.37$ | 0.60 |

